# Association of Cerebral Microbleeds and Risk of Stroke and Mortality in Posterior Circulation Cerebral Infarction

**DOI:** 10.1101/2024.07.25.24311035

**Authors:** Yajuan Wang, Xiaoyan Sun, Shasha Wu, Jianxiu Sun, Yuyuan Yang, Moxin Luan, Fei Yu, Jing Zhou, Xiaosa Chi, Xueping Zheng

## Abstract

**Objective:** This study was investigated to determine whether CMBs were associated with the risk of recurrent stroke or all-cause death in patients with acute posterior circulation cerebral infarction.

**Methods:** A retrospective analysis was conducted on 323 patients with acute posterior circulation cerebral infarction who aged ≥ 45 years and were hospitalized at Qingdao University Affiliated Hospital from January 1, 2016 to December 31, 2020. Patients were divided into different CMBs groups according to the presence, number and distribution of CMBs. Occurrence of stroke and death was recorded during follow-up. We drew Kaplan Meier survival curves and constructed Cox proportional hazards regression models based on different CMBs groups and clinical outcomes.

**Results:** A total of 323 patients were enrolled in our study, and 138 (42.72%) had CMBs. During a median follow-up of 1357 days, 87 (26.94%) experienced recurrent stroke or death. ≥5 CMBs (HR 1.723; 95% CI 1.021-2.907; P=0.041) and lobar CMBs (HR 2.312; 95% CI 1.204-4.441; P=0.012) were independent predictors associated with the composite risk of recurrent stroke and all-cause death. All CMBs statuses were not significantly correlated with the risk of recurrent stroke. The presence of CMBs (HR 3.358; 95% CI 1.259-8.954; P=0.015), ≥ 5 CMBs (HR 5.290; 95% CI 1.599-17.499; P=0.006) and deep CMBs (HR 3.265; 95% CI 1.003-10.628; P=0.049) were all independent factors associated with all-cause death.

**Conclusions:** In patients with acute posterior circulation cerebral infarction, ≥5 CMBs and lobar CMBs may increase the risk of poor clinical outcome (the composite of recurrent stroke and all-cause death). Furthermore, the presence CMBs, ≥ 5 CMBs and deep CMBs all independently may increase the risk of all-cause death.

## 1. Introduce

Cerebral microbleeds (CMBs) were subclinical lesions in the brain parenchyma caused by small vessel lesions, characterized by deposition of hemosiderin.^1^ As imaging markers of cerebral small vessel disease (CSVD), they could be detected by susceptibility-weighted imaging (SWI) of magnetic resonance imaging (MRI).^1^ There was a close correlation between CMBs and the risk of stroke and death in previous studies.^2,3^However, the prognostic impact of CMBs on the population with ischemic stroke was controversial.

A prospective cohort study suggested that the presence and number of CMBs were not significantly associated with risk of recurrent ischemic stroke and mortality in patients with acute ischemic stroke, but CMBs ≥10 were independent predictors of intracranial hemorrhage (ICH).^4^ In patients with ischemic stroke and atrial fibrillation, the presence of CMBs was associated with an increased relative risk of ICH and ischemic stroke, but it did not significantly increase the risk of vascular death.^5^ In non-cardiogenic ischemic stroke patients treated with antiplatelet therapy, CMBs ≥ 10 pronouncedly increased the risk of recurrent stroke.^6^ In anterior circulation acute ischemic stroke patients treated by endovascular thrombectomy, there was no significant association between the presence of CMBs and the risk of ICH.^7^ However, little was known on the relationship between CMBs and the clinic outcomes of posterior circulation stroke. 20-30% of ischemic stroke located in the posterior circulation, and small artery occlusion was most common in the TOAST etiology classification of posterior circulation stroke,^8^ indicating that cerebral small vessel lesions had a greater impact on posterior circulation stroke.

Therefore, in this study, we aimed to investigate whether CMBs were associated with the risk of stroke recurrence or all-cause death in patients with posterior circulation cerebral infarction.

## 2. Methods

### 2.1 Standard Protocol Approvals, Registrations, and Patient Consents

The ethics committee of the Affiliated Hospital of Qingdao University approved this study (NO. QYFYWZLL27329). Written informed consent of participants was waived because of a retrospective design and observational nature of this study.

### 2.2 Study design and participants

This was a retrospective and observational study in patients with acute posterior circulation cerebral infarction who underwent SWI scans and were retrospectively registered in the Affiliated Hospital of Qingdao University from January 1, 2016 to December 31, 2020. 385 patients with acute posterior circulation cerebral infarction, who aged ≥45 years and were confirmed through brain MRI scans within 48 hours following the onset of symptoms, were consecutively recruited. The exclusion criteria were as follows: (1) severe heart, liver, lung, or kidney failure, severe anemia, abnormal coagulation function, or advanced malignant tumors; (2) hemorrhagic stroke (intracerebral or subarachnoid hemorrhage); (3) incomplete clinical and examination data. we excluded 25 patients according to the exclusion criteria, 37 patients were lost to follow-up. Finally, a total of 323 participants were included for analysis.

### 2.3 Clinical background

We collected the following clinical data for all enrolled patients from inpatient database: age, sex, ever smoker, alcohol drinking, hypertension, diabetes, ischemic heart disease, remote stroke or transient ischemic attack (TIA), triglycerides (TG) and low-density lipoprotein cholesterol (LDL-C).

### 2.4 Brain MRI and Defining Criteria for CMBs Events

All patients were scanned using a 3.0T MRI scanner within 48 hours of admission. CMBs were detected on SWI. A CMB was defined as a circular or oval low-intensity lesion within the parenchyma, and measured approximately 2–10 mm on SWI.^1^ We described CMBs according to the Microbleed Anatomical Rating Scale (MARS).^9^ Patients were categorized into two groups (no CMBs and CMBs) based on the presence or absence of CMBs, three groups (no CMBs, 1–4 CMBs, and ≥5 CMBs) based on the number of CMBs or four groups (no CMBs, lobar CMBs, deep CMBs, and mixed CMBs) based on the distribution of CMBs. Lobar CMBs located in supratentorial strictly lobar (frontal, parietal, temporal, occipital, insula) or superficial cerebellum (gray matter, vermis). Deep CMBs located in deep regions, which included basal ganglia, thalamus, internal capsule, external capsule, corpus callosum, deep and periventricular white matter (DPWM), brainstem, and non-superficial cerebellum. Mixed CMBs simultaneously located in lobar and deep regions. Two trained neurologists (YY and ML), who were absolutely blinded to the clinical information, independently evaluated the number and distribution of CMBs on SWI images, along with the presence of white matter hyperintensities (WMH).They also evaluated the images of brain magnetic resonance angiography (MRA) or head-neck computer tomography angiography (CTA) to classify carotid stenosis or intracranial artery stenosis according to the North American Symptomatic Carotid Endarterectomy Trial (NASCET) or the Warfarin-Aspirin Symptomatic Intracranial Disease Study (WASID).^10, 11^ In cases of initial disagreement, the final number was reached through consensus.

### 2.5 Clinical Outcomes

The primary outcome was a composite of recurrent stroke and all-cause death, while recurrent stroke and death were independently considered as other outcomes. Recurrent stroke was defined as the rapid onset of new focal neurological deficits or the rapid deterioration of original symptoms and signs of focal neurological deficits, accompanied by clinical or imaging evidence of stroke, including ischemic stroke and ICH (it was excluded if ICH was directly related to thrombolysis). Follow-up started from the date of diagnosis with cranial MRI. All the patients were followed up by telephone or outpatient visits until recurrence stroke, death, refusal of further participation, loss to follow-up, or end of follow-up (May 31, 2022), whichever occurred first.

### 2.6 Statistical analysis

Variables were represented by mean (SD), median (IQR) or n (%) where applicable. The differences between groups at baseline were compared with the independent sample t test, the χ² test or Fisher’s exact test. We draw Kaplan Meier survival curves to estimate cumulative risk of clinical outcomes in patients based on different CMBs groups, and used the log-rank test to compare groups. We used Cox proportional hazard analysis to estimate hazard ratios (HRs) and 95% confidence intervals (CIs) for the occurrence of clinical outcomes of each CMBs group, using the no CMBs group as a reference. Three models were used: (1) an unadjusted model, (2) a model adjusted for age and sex, and (3) a model adjusted for age, sex, ever smoker, alcohol drinking, hypertension, diabetes, ischemic heart disease, remote stroke or TIA, TG, LDL-C, WMH, carotid stenosis or intracranial artery stenosis. We used SPSS 25.0 (Chicago, IL) and GraphPad Prism (version 9.0; GraphPad, Inc., La Jolla, CA, USA) to perform the statistical analyses. P < 0.05 was considered to be significant.

## 3. Results

### 3.1 Baseline characteristics of the study participants

A total of 385 patients with acute posterior circulation cerebral infarction were consecutively screened in our cohort study according to the inclusion criteria, and 25 out of 385 (6.49%) patients were excluded for various reasons (Figure 1). During a median follow-up of 1357 (IQR1242-1471) days, 37 patients were lost to follow-up. Finally, a total of 323 participants were included for analysis (Figure 1).

**Figure 1.**
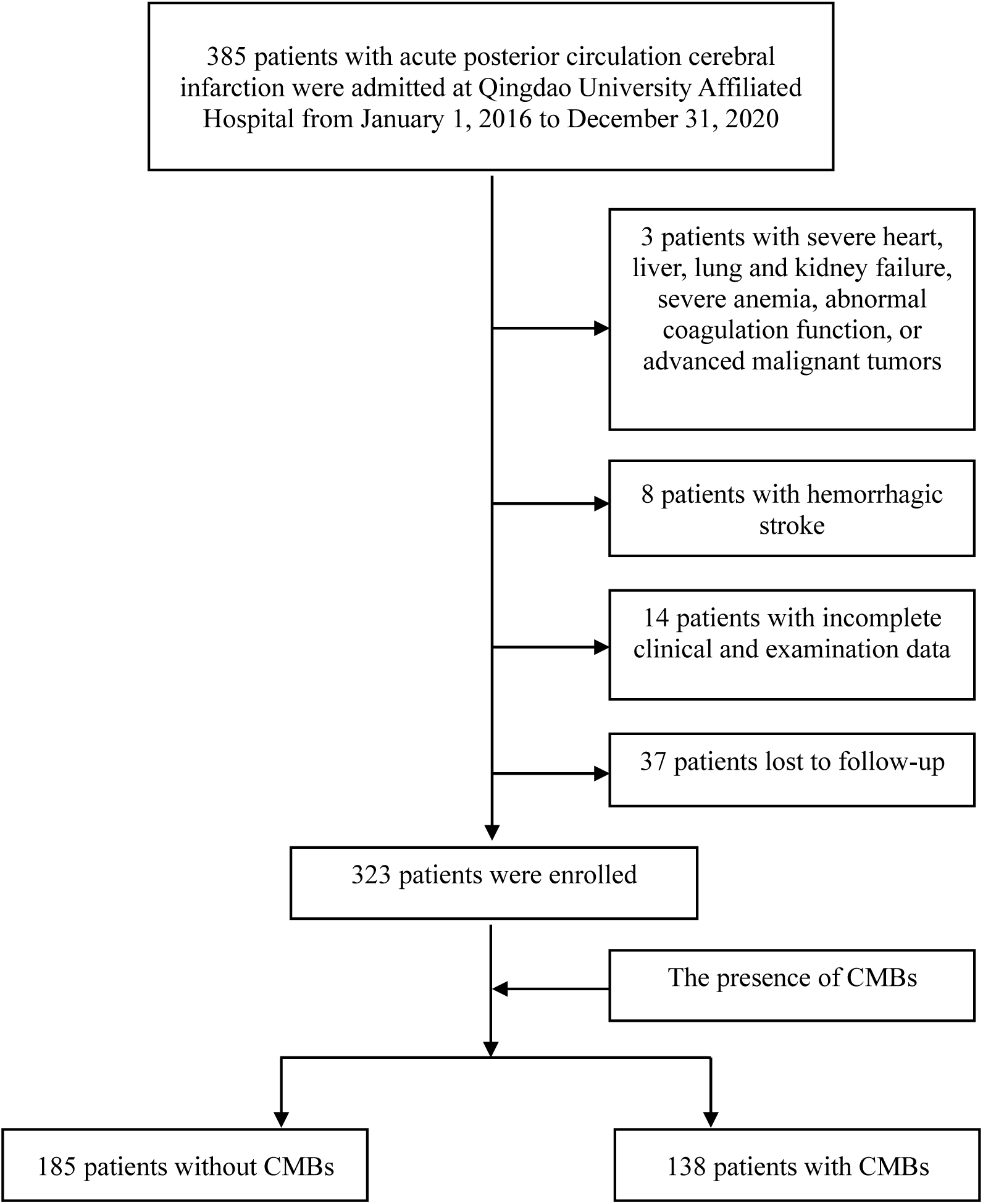
Study flow chart.

The clinical and neuroimaging characteristics of the study population were shown in **Table 1**. The mean age of the study population at baseline was 66.12 years, 56.35% were ≥65 years, and 69.97% were men. Overall, 71.21% had a past history of hypertension and 27.55% had a past history of remote stroke or TIA. In addition, 42.72% (138/323) had presence of CMBs, of which 49.28% (68/138) had 1-4 CMBs, and 50.72% (70/138) had ≥5 CMBs. Furthermore, 22.46% (31/138), 43.48% (60/183) and 34.06% (47/138) of patients with CMBs were respectively lobar, deep and mixed in distribution. Moreover, 94.43% (305/323) of the population regularly took antiplatelet drugs in hospital and after discharge.

**Table 1.** Clinical characteristics and outcomes of the study population (n=323)

| Characteristics at baseline | Values |
| --- | --- |
| **Age** |  |
| y, mean (SD) | 66.12(10.12) |
| ≥65, n (%) | 182(56.35) |
| **Sex** |  |
| Male, n (%) | 226(69.97) |
| **Ever smoker**, n (%) | 118(36.53) |
| **Alcohol drinking**, n (%) | 102(31.58) |
| **Comorbidities** |  |
| Hypertension, n (%) | 230(71.21) |
| Diabetes, n (%) | 142(43.96) |
| Ischemic heart disease, n (%) | 71(21.98) |
| Remote stroke or TIA (>6 months prior), n (%) | 89(27.55) |
| **Laboratory examinations** |  |
| TG, mmol/L, n (%) |  |
| ≥1.7 | 93(28.79) |
| LDL-C, mmol/L, n (%) |  |
| ≥3.4 | 43(13.31) |
| **Neuroimaging** |  |
| CMBs, n (%) |  |
| Presence of CMBs | 138(42.72) |
| 1-4 CMBs | 68(21.05) |
| ≥5 CMBs | 70(21.67) |
| Lobar CMBs | 31(9.60) |
| Deep CMBs | 60(18.56) |
| Mixed CMBs | 47(14.56) |
| WMH, n (%) | 246(76.16) |
| Carotid stenosis or intracranial artery stenosis, n (%) |  |
| <30% | 71(21.98) |
| 30%-49% | 121(37.46) |
| ≥50% | 131(40.56) |
| **Therapy** |  |
| Regular antiplatelet therapy (in hospital and after discharge), n (%) | 305(94.43) |
| Regular statin therapy (in hospital and after discharge), n (%) | 310(95.98) |
| **Outcome** |  |
| Follow-up time, d, median (IQR) | 1357(1242-1471) |
| Composite of recurrent stroke and all-cause death, n (%) | 87(26.94) |
| Recurrent stroke, n (%) | 65(20.12) |
| Ischemic stroke, n (%) | 60(18.58) |
| ICH, n (%) | 5(1.54) |
| All-cause death, n (%) | 22(6.81) |

Abbreviations: TIA, transient ischemic attack; TG, triglyceride; LDL-C, low-density lipoprotein cholesterol; CMBs, cerebral microbleeds; WMH, white matter hyperintensities; ICH, intracranial hemorrhage. Data are presented as mean (SD), median (IQR) or n (%).

A comparison of clinical and neuroimaging characteristics of patients with and without CMBs was shown in **Table 2**. Compared with patients without CMBs, patients with CMBs were older (p=0.003), had a higher prevalence of ≥65 years (P=0.011) and were more likely to have a history of remote stroke or TIA (P =0.001). Patients with CMBs also tended to have WMH (P=0.009).

**Table 2.**
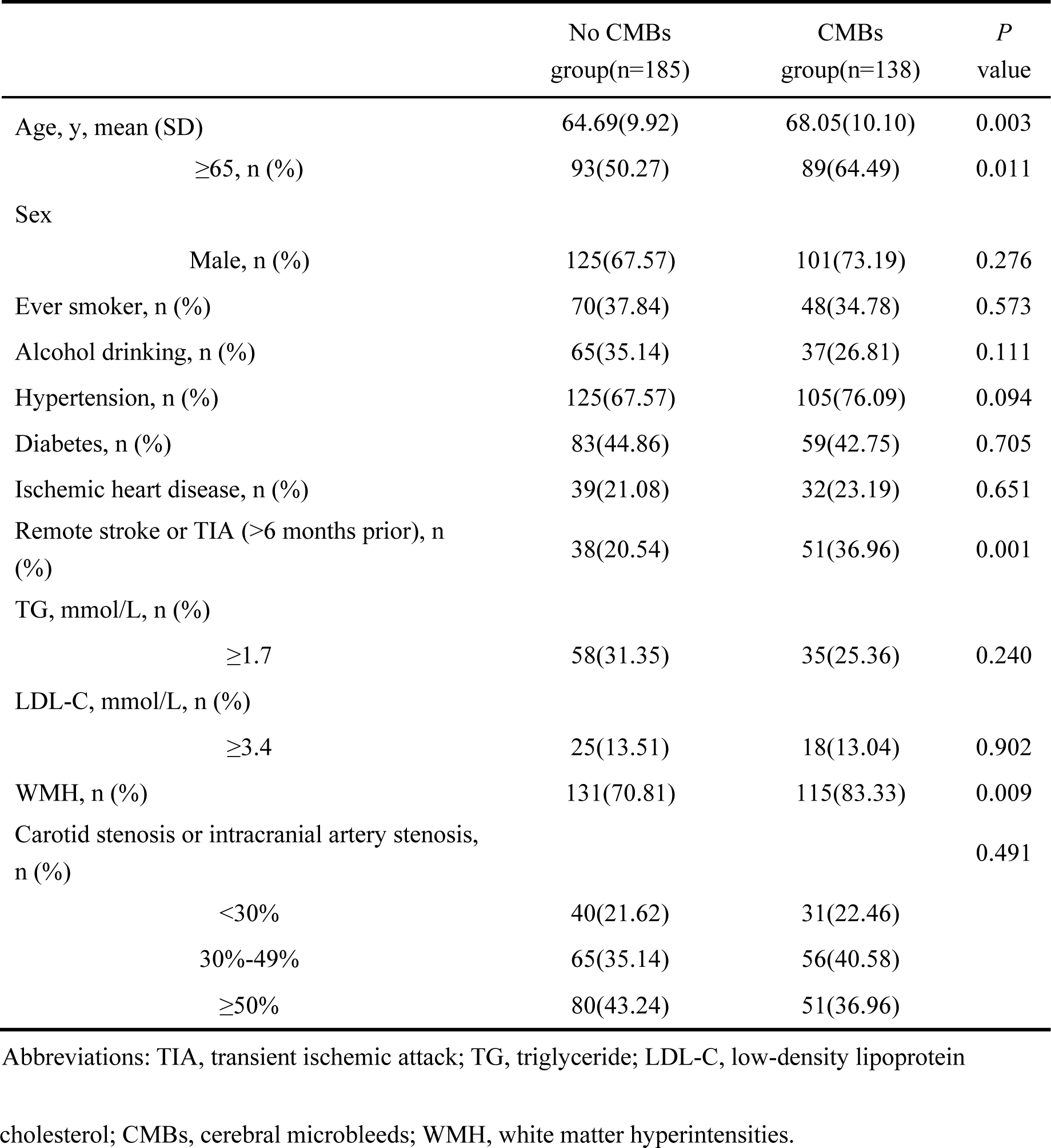
Baseline characteristics of the study population based on the presence of CMBs.

### 3.2 Clinical outcomes

Overall, 87 patients in the study population experienced the primary outcome (a composite of recurrent stroke and all-cause death) during the follow-up. 65 patients experienced recurrent stroke and 22 patients died (**Table 1**).

#### 3.2.1 The presence of CMBs and clinical outcomes

Compared with patients without CMBs, patients with CMBs at baseline were at increased risk of the primary outcome (HR 1.653; 95% CI 1.084-2.519; P=0.019) (**Figure 2**A). However, there were no significant differences in the primary outcome between no CMBs group and CMBs group after adjusting for multivariate variables (P > 0.05) (**Figure 2**D). The multivariate Cox proportional hazards regression analysis revealed that LDL-C≥3.4mmol/L (HR 1.908; 95% CI 1.052–3.461; P = 0.033) and WMH (HR 1.900; 95% CI 1.034–3.490; P = 0.039) were all significantly related to the occurrence of the primary outcome (**Figure 2**D). In addition, the presence of CMBs was not significantly associated with the risk of recurrent stroke compared with no CMBs (P > 0.05) (**Table 4**). However, compared with no CMBs, the presence of CMBs (HR 3.358; 95% CI 1.259-8.954; P = 0.015) were independently associated with the risk of all-cause death after adjusting for multivariate variables (**Table 4**).

**Figure 2.**
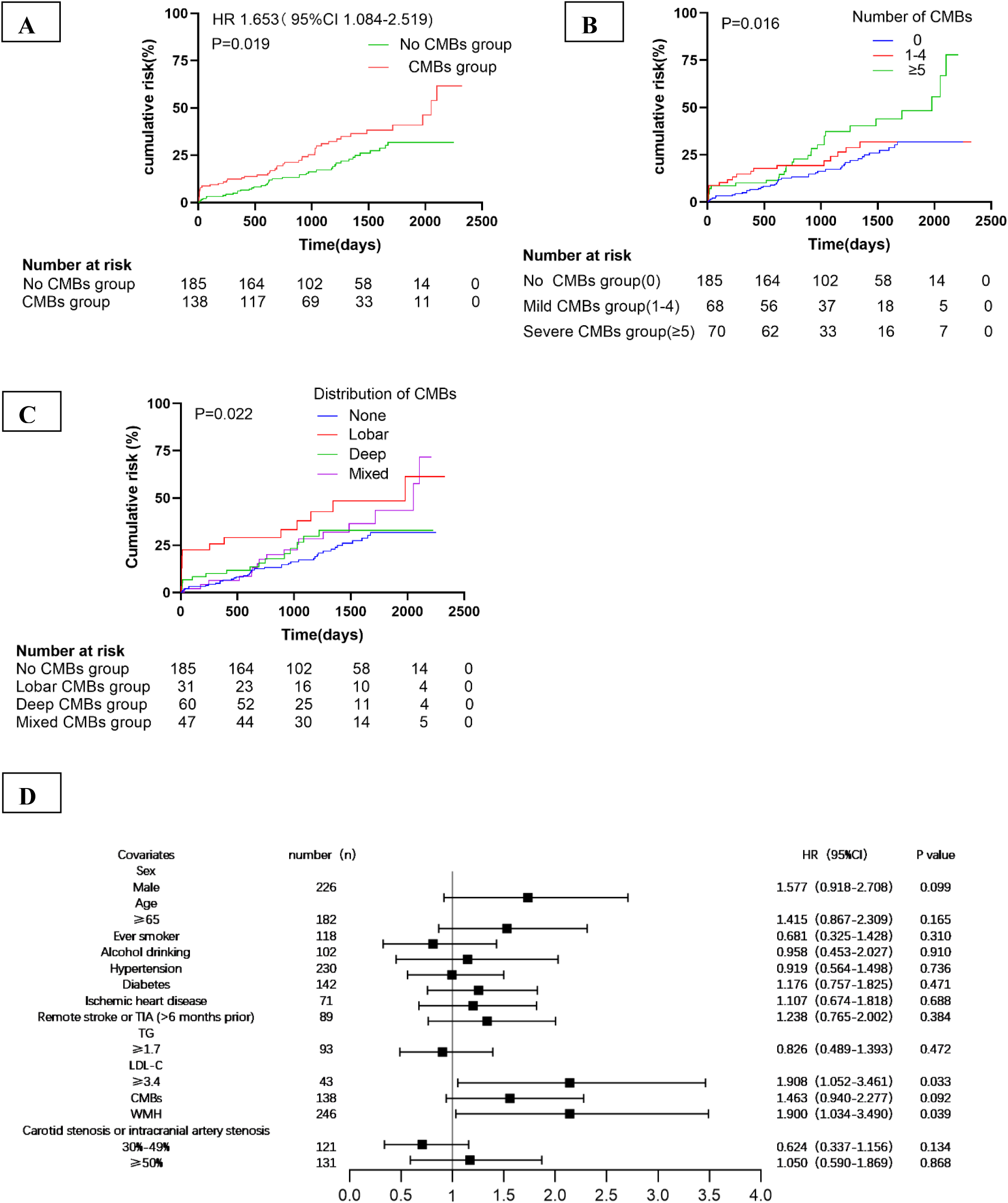
Cumulative incidence of the primary outcome and multivariate cox regression analysis of the primary outcome. **A**, Cumulative incidence of the primary outcome—a composite of recurrence stroke and all-cause death among the two groups (no CMBs group and CMBs group). **B**, Cumulative incidence of the primary outcome among the three groups (no CMBs group, 1–4 CMBs group and ≥5 CMBs group). **C**, Cumulative incidence of the primary outcome among the four groups (no CMBs group, lobar CMBs group, deep CMBs group and mixed CMBs group). CMBs indicate cerebral microbleeds; HR indicates hazard ratio. **D**, The forest plot shows cox proportional hazards regression analysis of the primary outcome—a composite of recurrence stroke and all-cause death. **Abbreviation**: HR, hazard ratio; CI, confidence interval; TIA, transient ischemic attack; TG, triglyceride; LDL-C, low-density lipoprotein cholesterol; CMBs, cerebral microbleeds; WMH, white matter hyperintensities.

#### 3.2.2 The number of CMBs and clinical Outcomes

Compared with patients without CMBs, patients with 1-4 CMBs had a similar rate of the primary outcome after adjusting for multivariate variables (P = 0.092) (**Figure 2**B and **Table 3**). However, the rate of the primary outcome was significantly higher in patients with ≥5 CMBs than in those without CMBs (HR 2.002; 95% CI 1.237-3.241; P = 0.005). The multivariate Cox proportional hazards regression analysis revealed that ≥5 CMBs (HR 1.723; 95% CI 1.021-2.907; P = 0.041) were still significantly associated with the occurrence of the primary outcome (**Table 3**). In addition, compared with no CMBs, 1-4 CMBs and ≥5 CMBs were not associated with the risk of recurrent stroke (P > 0.05) (**Table 4**). Nevertheless, the Cox proportional hazards regression analysis showed that, compared with no CMBs, ≥5 CMBs (HR 5.290; 95% CI 1.599-17.499; P = 0.006) were independently associated with the risk of all-cause death after adjusting for multivariate variables (**Table 4**).

**Table 3.** Cox regression analyses on the risk of primary outcome according to the presence, number and distribution of baseline CMBs.

|  | n (%) | Unadjusted HR<br>(95% CI) | P<br>value | Age and Sex<br>Adjusted HR (95%<br>CI) | P<br>value | Multivariate<br>Adjusted HR (95%<br>CI) * | P value |
| --- | --- | --- | --- | --- | --- | --- | --- |
| No CMBs (n=185) | 41(22.16) | 1(ref) | - | 1(ref) | - | 1(ref) | - |
| CMBs presence<br>(n=138) | 46(33.33) | 1.653(1.084-2.519) | 0.019 | 1.595(1.045-2.436) | 0.031 | 1.463(0.940-2.277) | 0.092 |
| **Number** |  |  |  |  |  |  |  |
| 1-4 CMBs(n=68) | 18(26.47) | 1.301(0.748-2.266) | 0.352 | 1.313(0.751-2.294) | 0.339 | 1.220(0.692-2.149) | 0.492 |
| $\geq 5$ CMBs(n=70) | 28(40.00) | 2.002(1.237-3.241) | 0.005 | 1.860(1.140-3.035) | 0.013 | 1.723(1.021-2.907) | 0.041 |
| **Distribution** |  |  |  |  |  |  |  |
| Lobar CMBs(n=31) | 14(45.16) | 2.462(1.342-4.519) | 0.004 | 2.224(1.199-4.126) | 0.011 | 2.312(1.204-4.441) | 0.012 |
| Deep CMBs(n=60) | 15(25.00) | 1.308(0.723-2.366) | 0.374 | 1.340(0.739-2.431) | 0.335 | 1.117(0.606-2.059) | 0.723 |
| Mixed CMBs(n=47) | 17(36.17) | 1.588(0.902-2.797) | 0.109 | 1.509(0.854-2.665) | 0.156 | 1.412(0.767-2.598) | 0.268 |

Estimates represent hazard ratios (95% CI) for CMBs statuses. No CMBs group was the reference.
\* Adjusted for age, sex, ever smoker, alcohol drinking, hypertension, diabetes, ischemic heart disease, remote stroke or TIA, TG, LDL-C, WMH, carotid stenosis or intracranial artery stenosis. Abbreviations: HR, hazard ratio; CI, confidence interval; CMBs, cerebral microbleeds; TIA, transient ischemic attack; TG, triglyceride; LDL-C, low-density lipoprotein cholesterol; WMH, white matter hyperintensities.

#### 3.2.3 The distribution of CMBs and clinical Outcomes

There was a significant difference in the primary outcome between the four groups (no CMBs, lobar CMBs, deep CMBs, and mixed CMBs) (P = 0.022) (**Figure 2**C). Compared with patients without CMBs, patients with mixed and deep CMBs had similar HRs for the primary outcome (P > 0.05), but patients with lobar CMBs (HR 2.462; 95% CI 1.342-4.519; P = 0.004) had a higher risk of the primary outcome (**Table 3**). The multivariate Cox proportional hazards regression analysis showed that lobar CMBs (HR 2.312; 95% CI 1.204-4.441; P = 0.012) were still significantly associated with the occurrence of the primary outcome (**Table 3**). In addition, compared with no CMBs, only lobar CMBs (HR 2.194; 95% CI 1.081-4.453; P = 0.030) had a higher risk of recurrent stroke (**Table 3**). However, the multivariate Cox proportional hazards regression analysis showed that lobar CMBs were no independently associated with the risk of recurrent stroke (P = 0.054) (**Table 4**). Compared with no CMBs, mixed CMBs and deep CMBs had similar HRs for the risk of all-cause death (P > 0.05), lobar CMBs (HR 3.559; 95% CI 1.069-11.849; P = 0.039) had a higher risk of all-cause death (**Table 4**). However, the Cox proportional hazards regression analysis showed that, deep CMBs (HR 3.265; 95% CI 1.003-10.628; P = 0.049), but not lobar CMBs (HR 2.105; 95% CI 0.988-4.485; P = 0.054), were independently associated with the risk of all-cause death after adjusting for multivariate variables (**Table 4**).

**Table 4.** Cox regression analyses on the risk of other outcomes according to the presence, number and distribution of baseline CMBs.

|  | n(%) | Unadjusted HR<br>(95% CI) | P<br>value | Age and Sex Adjusted<br>HR (95% CI) | P<br>value | Multivariate Adjusted<br>HR (95% CI) * | P<br>value |
| --- | --- | --- | --- | --- | --- | --- | --- |
| **Recurrent stroke** |  |  |  |  |  |  |  |
| No CMBs (n=185) | 33(17.84) | 1(ref) | - | 1(ref) | - | 1(ref) | - |
| CMBs<br>presence(n=138) | 32(23.19) | 1.423(0.875-2.316) | 0.155 | 1.391(0.853-2.269) | 0.186 | 1.237(0.742-2.060) | 0.414 |
| Number |  |  |  |  |  |  |  |
| 1-4 CMBs(n=68) | 13(19.12) | 1.161(0.611-2.207) | 0.648 | 1.197(0.627-2.283) | 0.586 | 1.083(0.561-2.089) | 0.812 |
| ≥5 CMBs(n=70) | 19(27.14) | 1.685(0.957-2.967) | 0.071 | 1.572(0.885-2.792) | 0.123 | 1.390(0.757-2.550) | 0.288 |
| Distribution |  |  |  |  |  |  |  |
| Lobar CMBs(n=31) | 10(32.26) | 2.194(1.081-4.453) | 0.030 | 2.048(0.995-4.216) | 0.052 | 2.105(0.988-4.485) | 0.054 |
| Deep CMBs(n=60) | 9(15.00) | 0.965(0.461-2.020) | 0.925 | 0.995(0.474-2.086) | 0.989 | 0.834(0.390-1.783) | 0.640 |
| Mixed<br>CMBs(n=47) | 13(27.66) | 1.508(0.793-2.867) | 0.211 | 1.451(0.760-2.768) | 0.259 | 1.257(0.632-2.502) | 0.514 |
| **All-cause death** |  |  |  |  |  |  |  |
| No CMBs (n=185) | 8(4.32) | 1(ref) | - | 1(ref) | - | 1(ref) | - |
| CMBs<br>presence(n=138) | 14(10.14) | 2.603(1.091-6.211) | 0.031 | 2.432(1.017-5.817) | 0.046 | 3.358(1.259-8.954) | 0.015 |
| Number |  |  |  |  |  |  |  |
| 1-4 CMBs(n=68) | 5(7.35) | 1.882(0.616-5.757) | 0.267 | 1.780(0.576-5.503) | 0.317 | 2.356(0.717-7.735) | 0.158 |
| ≥5 CMBs(n=70) | 9(12.86) | 3.313(1.275-8.608) | 0.014 | 3.078(1.162-8.154) | 0.024 | 5.290(1.599-17.499) | 0.006 |
| Distribution |  |  |  |  |  |  |  |
| Lobar CMBs(n=31) | 4(12.90) | 3.559(1.069-11.849) | 0.039 | 2.992(0.886-10.109) | 0.078 | 3.670(0.938-14.364) | 0.062 |
| Deep CMBs(n=60) | 6(10.00) | 2.764(0.956-7.986) | 0.060 | 2.815(0.963-8.225) | 0.059 | 3.265(1.003-10.628) | 0.049 |
| Mixed CMBs(n=47) | 4(8.51) | 1.920(0.578-6.380) | 0.287 | 1.748(0.524-5.830) | 0.364 | 3.087(0.733-12.996) | 0.124 |

Estimates represent hazard ratios (95% CI) for CMBs statuses. No CMBs group was the reference.
\* Adjusted for age, sex, ever smoker, alcohol drinking, hypertension, diabetes, ischemic heart disease, remote stroke or TIA, TG, LDL-C, WMH, carotid stenosis or intracranial artery stenosis. Abbreviations: HR, hazard ratio; CI, confidence interval; CMBs, cerebral microbleeds; TIA, transient ischemic attack; TG, triglyceride; LDL-C, low-density lipoprotein cholesterol; WMH, white matter hyperintensities.

## 4. Discussion

To our knowledge, no previous studies reported on the association of CMBs with long-term clinical outcomes in patients with acute posterior circulation cerebral infarction. In our present study, we demonstrated that ≥5 CMBs and lobar CMBs, but not the presence of CMBs (CMBs ≥1), were significantly associated with the risk of the primary outcome (a composite of recurrent stroke and all-cause death) in patients with acute posterior circulation cerebral infarction. In addition, LDL-C ≥3.4mmol/L and WMH were also significantly related to the occurrence of the primary outcome. However, all CMBs statuses were not associated with the risk of recurrent stroke. Furthermore, the presence of CMBs, ≥5 CMBs, and deep CMBs were independently associated with the risk of all-cause death.

In our retrospective observational cohort study, 42.72% of patients with acute posterior circulation cerebral infarction were noted to have CMBs. It was similar to the prevalence noted in previous study of ischemic stroke individuals (35–71%),^12^ but higher than the prevalence in patients with a first-ever anterior circulation ischemic stroke (23.4%).^13^ A multi-center MRI study demonstrated that, compared with anterior circulation ischemic stroke, small artery occlusion was more common in posterior circulation stroke,^8^ therefore, as a marker of CSVD, CMBs may be more common in posterior circulation stroke. Moreover, our patients were middle-aged and elderly people, and some of them were suffered from recurrent stroke, which may contribute to a higher prevalence of CMBs in our patients. The ratios of deep CMBs and mixed CMBs were higher than those of lobar CMBs in our participants. The pathogenesis of CMBs is not yet clear. It is mainly believed to be related to hypertensive vascular damage and cerebral amyloid angiopathy (CAA) based on anatomical distribution. Hypertension-related CMBs located in the deep and infratentorial regions, because these regions may be mainly supplied by deep perforating branches, which were prone to atherosclerosis due to hypertension, vascular endothelial damage led to blood extravasation.^14^ CAA-related CMBs located in the lobar regions, because these regions may be mainly supplied by cortical and leptomeningeal vessels, on where β-amyloid protein may be more likely to deposit and activate immune response, causing damage to brain parenchyma and blood vessels by releasing neurotoxic factors and leading to CMBs.^14^ CMBs located in the mixed regions may had both of the above mechanisms simultaneously. In our study population, the incidence of hypertension in the CMBs group was as high as 76.09%, which may lead to a higher prevalence of deep and mixed CMBs. Different from the anatomical distribution of CMBs in Western populations, our result was in line with an individual participant meta-analysis, which affirmed that Eastern populations had a higher prevalence of deep or mixed CMBs compared with Western populations.^15^ Our study also found that patients with CMBs were older and more likely to have a history of remote stroke or TIA, and to have WMH than those without CMBs. As shown in previous studies, the overall prevalence of CMBs increased with age,^16^ CMBs were common in stroke patients and more prevalent among patients with recurrent stroke.^12^ As neuroimaging markers of CSVD, both CMBs and WMH could reflect the presence and severity of microvascular damage, they often coexisted in patients with cerebrovascular diseases, especially in middle-aged and elderly stroke patients.

We found no statistically significant differences in the risk of primary outcome between no CMBs group and CMBs group after adjusting for multiple covariates. Only LDL-C ≥3.4mmol/L and WMH were significantly associated with the risk of the primary outcome. Statin therapy or a more intensive statin regimen produced a 21% proportional reduction in the risk of major vascular events per 1.0 mmol/L reduction in LDL-C.^17^ Both European Stroke Organisation and American Stroke Association suggested the ischemic stroke or TIA second prevention guideline with a recommendation of target LDL-C <1.8 mmol/L. ^18, 19^ However, lower LDL-C level (≤1.76 mmol/L) was related with higher CMB development,^20^ a prospective study observed a significant association between lower LDL-C level and higher risk of ICH when LDL-C was <1.8mmol/L.^21^ There was no consensus on target for LDL-C level in patients with posterior circulation infarction. Atorvastatin therapy may reduce the occurrence of new deep CMBs,^20^ pravastatin treatment significantly reduced the risk of recurrent stroke among patients with posterior circulation stroke,^22^ which may benefit for posterior circulation stroke patients with CMBs. But the optimal LDL-C level and intensive dose of statin are not yet clear, further clinic trials are warranted in the near future. A prospective cohort study reported that the presence of CMBs was associated with lower mean fractional anisotropy and higher mean diffusivity, the radial diffusivity of the CMBs group was higher,^23^ indicating that microstructural destruction of brain white matter existed in CMBs. Previous studies suggested that WMH increased the risk of CMBs,^16, 24^ A high microbleed burden (≥5) was independently associated with a severe burden of subcortical and periventricular WMH.^25^ A previous study reported that WMH volume, type, and shape, especially confluent WMH, were independently associated with long-term risk of mortality and ischemic stroke, because this type of WMH was likely to represent more severe brain parenchymal damage.^26^ There were high prevalence of CMBs and WMH in our patients, they may affect each other and develop together, therefore, the coexistence of CMBs and WMH was more likely to increase the risk of poor prognosis in stroke patients. Further analysis revealed that ≥5 CMBs were significantly related to the occurrence of the primary outcome. An explanation for this finding may be that ≥5 CMBs represented more severe and extensive cerebral small vessel lesions, may lead to poor outcomes. Our study also showed that lobar CMBs were significantly related to the occurrence of the primary outcome, which was consistent with a previous study, the study confirmed that lobar CMBs clearly increased the risk of recurrent stroke and all-cause mortality in participants with embolic stroke of undetermined source.^27^ Therefore, it is essential to consider the number and distribution of CMBs while we develop strategies for treatment of posterior circulation infarction.

In our study, we observed that there were no statistically significant differences between all CMBs statuses and the risk of recurrent stroke. So far, there was no consensus on the relationship between CMBs and the risk of recurrent stroke. A study recruited 1003 Chinese patients with ischemic stroke during a mean follow-up of 37 months and found that, neither CMBs distribution nor burden was associated with the risk of recurrent ischemic stroke, but there was a significant correlation between ≥ 5 CMBs and an increased risk of ICH.^25^ Evidence that the absolute risk of future ischemic stroke remained significantly higher than that of future ICH in patients with recent ischemic stroke or TIA, was found in a recent meta-analysis of clinical studies.^28^ The study also found that the number of CMBs was related to the risk of recurrent stroke (CMBs≥1, HR 1.35, 95% CI 1.20-1.50), especially ICH (CMBs=1, HR 1.87, 95% Cl 1.23-2.84; CMBs=2-4, HR 1.89, 95% Cl 1.22-2.93; CMBs ≥ 5, HR 4.55, 95% Cl 3.08-6.72), however, the distribution of CMBs had little effect on the risk of recurrent ischemic stroke.^28^ In our study, we also found that the risk of recurrent ischemic stroke was significantly higher than the risk of ICH, but we did not find a significant impact of CMBs on the future stroke risk of posterior circulation cerebral infarction. An explanation for this finding may be that the distribution of cerebral infarction in our participants were different from other studies. A previous study also supported different prognosis in stroke of different regions^29^. We observed that CMBs did not significantly increase the risk of recurrent stroke, a clinical study observed that the risk of ICH after intravenous thrombolysis in posterior circulation stroke was lower than that of anterior circulation stroke.^30^ Therefore, it is unwise to refuse antithrombotic therapy in patients with posterior circulation infarction combined with CMBs because of being overly concerned about the risk of future ICH.

We observed that the presence of CMBs was significantly associated with the risk of all-cause mortality in patients with acute posterior circulation cerebral infarction, which was in line with a previous report.^2^ An explanation for the association with mortality may be that CMBs reflected severe diffuse vascular pathology and frailty, as well as disease-associated vascular risk factors.^2^ Further analysis showed that the number of CMBs ≥5 was an independent risk factor for all-cause mortality. ≥5 CMBs may reflect more severe and extensive microvascular damage, leading to adverse outcomes. In the population with acute posterior circulation cerebral infarction, we found that lobar CMBs were associated with the risk of all-cause mortality on univariate analysis, but this did not reach statistical significance on multivariate analysis. On the contrary, our study suggested that deep CMBs were independently associated with the risk of all-cause mortality. Therefore, more aggressive management strategies are needed among posterior circulation infarction patients combined with CMBs.

There were some limitations in our study. First, it was a single-center and retrospective study, which could not represent the general population of acute posterior circulation cerebral infarction. Second, some patients with posterior circulation infarction, especially brainstem infarction, may be DWI-negative, and were not included. Moreover, patients with posterior circulation infarction who did not perform SWI scans for various reasons were also excluded, which may result in selection bias. Third, the participants of this study were middle-aged and elderly people, it was reported that the prevalence of CMBs increased along with age,^16^ and advanced age was independently associated with poor prognosis in patients with stroke.^4, 6^ However, we could not further analyze the groups of different age levels.

In conclusion, our findings suggest that CMBs were significantly associated with poor prognosis, especially all-cause mortality, in patients with acute posterior circulation infarction. Therefore, it seems proper to consider CMBs statuses while we investigate the risk of future poor clinical outcomes in patients with acute posterior circulation infarction. The preventive treatment studies of CMBs are required in the near future. In addition, future studies are needed on secondary prevention in posterior circulation infarction patients with CMBs, especially with a high burden of CMBs. The issue that how to balance the relationship between CMBs, LDL-C level and statins also should be conducted in future to improve the poor prognosis of posterior circulation infarction.

## Acknowledgments

We thank all the participants and the research staff for their contribution to the study.

## Data availability statement

All datesets are available free upon request to Prof Xueping Zheng.

## Author contributions

Y. Wang provided study design, collected data, did the statistical analysis and interpretation, wrote the manuscript, X. Sun, S. Wu, J. Sun did the statistical analysis and performed the literature search, Y. Yang and M. Luan collected data, F. Yu, J. Zhou, and X. Chi contributed to data management, X. Zheng designed the project conception, obtained funding, provided study supervision. All authors contributed to critical revision of the manuscript and approved the final manuscript for submission.

---

STROBE Statement—checklist of items that should be included in reports of observational studies

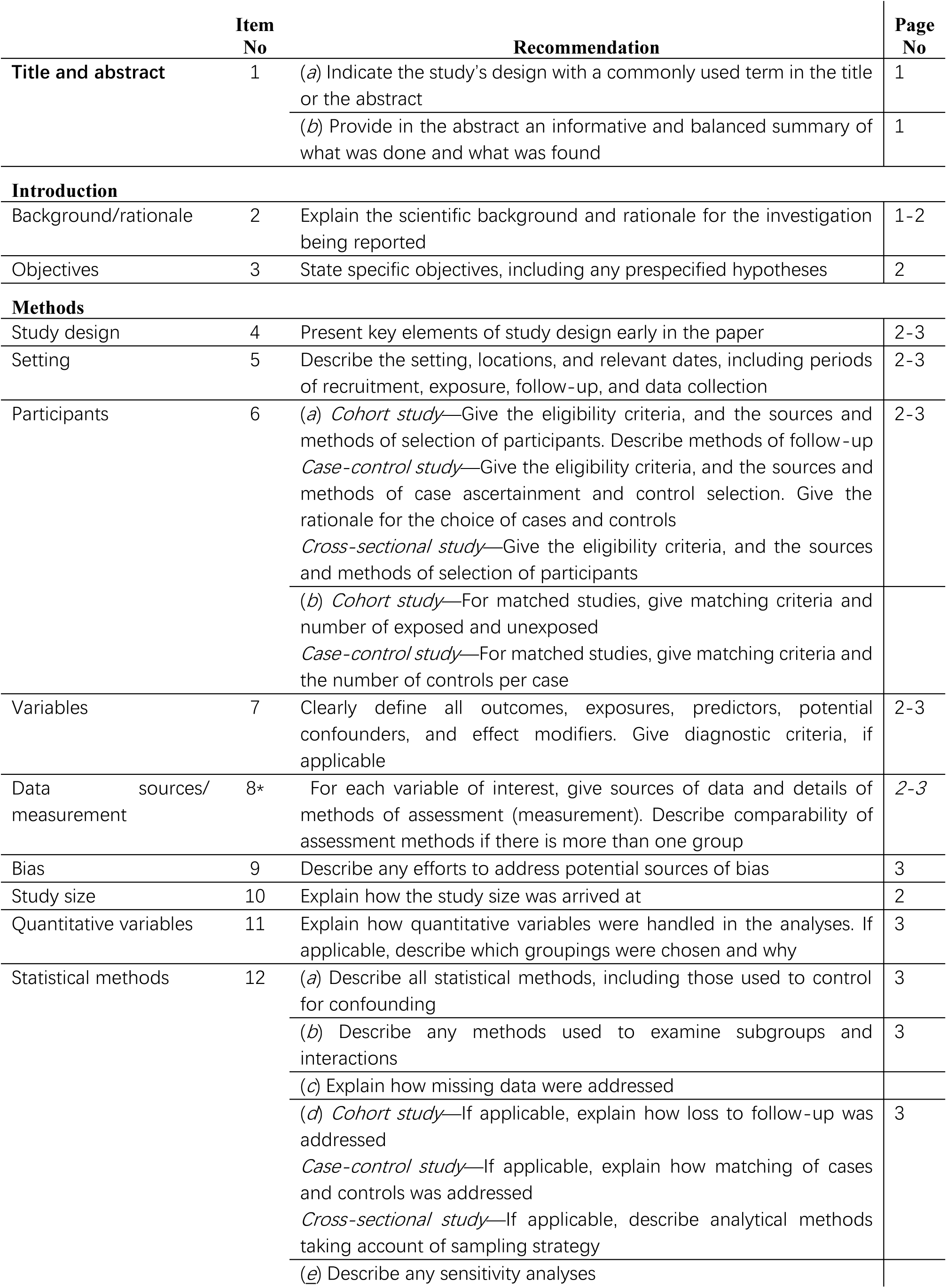
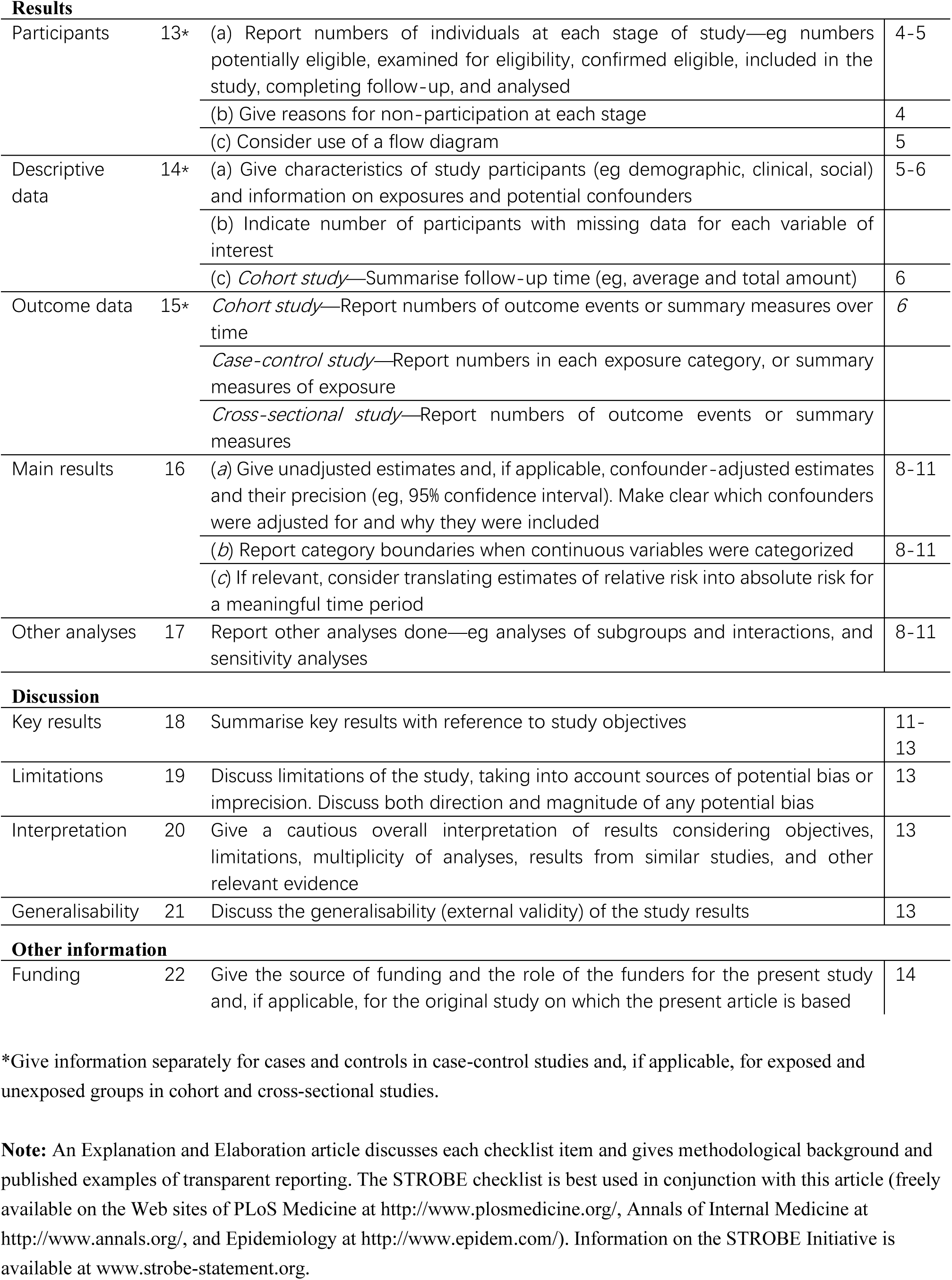

|  | Item No | Recommendation | Page No |
| --- | --- | --- | --- |
| Title and abstract | 1 | (a) Indicate the study's design with a commonly used term in the title or the abstract | 1 |
|  |  | (b) Provide in the abstract an informative and balanced summary of what was done and what was found | 1 |
| **Introduction** |  |  |  |
| Background/rationale | 2 | Explain the scientific background and rationale for the investigation being reported | 1-2 |
| Objectives | 3 | State specific objectives, including any prespecified hypotheses | 2 |
| **Methods** |  |  |  |
| Study design | 4 | Present key elements of study design early in the paper | 2-3 |
| Setting | 5 | Describe the setting, locations, and relevant dates, including periods of recruitment, exposure, follow-up, and data collection | 2-3 |
| Participants | 6 | (a) *Cohort study*—Give the eligibility criteria, and the sources and methods of selection of participants. Describe methods of follow-up<br>*Case-control study*—Give the eligibility criteria, and the sources and methods of case ascertainment and control selection. Give the rationale for the choice of cases and controls<br>*Cross-sectional study*—Give the eligibility criteria, and the sources and methods of selection of participants | 2-3 |
|  |  | (b) *Cohort study*—For matched studies, give matching criteria and number of exposed and unexposed<br>*Case-control study*—For matched studies, give matching criteria and the number of controls per case |  |
| Variables | 7 | Clearly define all outcomes, exposures, predictors, potential confounders, and effect modifiers. Give diagnostic criteria, if applicable | 2-3 |
| Data sources/<br>measurement | 8* | For each variable of interest, give sources of data and details of methods of assessment (measurement). Describe comparability of assessment methods if there is more than one group | 2-3 |
| Bias | 9 | Describe any efforts to address potential sources of bias | 3 |
| Study size | 10 | Explain how the study size was arrived at | 2 |
| Quantitative variables | 11 | Explain how quantitative variables were handled in the analyses. If applicable, describe which groupings were chosen and why | 3 |
| Statistical methods | 12 | (a) Describe all statistical methods, including those used to control for confounding | 3 |
|  |  | (b) Describe any methods used to examine subgroups and interactions | 3 |
|  |  | (c) Explain how missing data were addressed |  |
|  |  | (d) *Cohort study*—If applicable, explain how loss to follow-up was addressed<br>*Case-control study*—If applicable, explain how matching of cases and controls was addressed<br>*Cross-sectional study*—If applicable, describe analytical methods taking account of sampling strategy | 3 |
|  |  | (e) Describe any sensitivity analyses |  |
| **Results** |  |  |  |
| Participants | 13* | (a) Report numbers of individuals at each stage of study—eg numbers potentially eligible, examined for eligibility, confirmed eligible, included in the study, completing follow-up, and analysed | 4-5 |
|  |  | (b) Give reasons for non-participation at each stage | 4 |
|  |  | (c) Consider use of a flow diagram | 5 |
| Descriptive data | 14* | (a) Give characteristics of study participants (eg demographic, clinical, social) and information on exposures and potential confounders | 5-6 |
|  |  | (b) Indicate number of participants with missing data for each variable of interest |  |
|  |  | (c) *Cohort study*—Summarise follow-up time (eg, average and total amount) | 6 |
| Outcome data | 15* | *Cohort study*—Report numbers of outcome events or summary measures over time | 6 |
|  |  | *Case-control study*—Report numbers in each exposure category, or summary measures of exposure |  |
|  |  | *Cross-sectional study*—Report numbers of outcome events or summary measures |  |
| Main results | 16 | (a) Give unadjusted estimates and, if applicable, confounder-adjusted estimates and their precision (eg, 95% confidence interval). Make clear which confounders were adjusted for and why they were included | 8-11 |
|  |  | (b) Report category boundaries when continuous variables were categorized | 8-11 |
|  |  | (c) If relevant, consider translating estimates of relative risk into absolute risk for a meaningful time period |  |
| Other analyses | 17 | Report other analyses done—eg analyses of subgroups and interactions, and sensitivity analyses | 8-11 |
| **Discussion** |  |  |  |
| Key results | 18 | Summarise key results with reference to study objectives | 11-13 |
| Limitations | 19 | Discuss limitations of the study, taking into account sources of potential bias or imprecision. Discuss both direction and magnitude of any potential bias | 13 |
| Interpretation | 20 | Give a cautious overall interpretation of results considering objectives, limitations, multiplicity of analyses, results from similar studies, and other relevant evidence | 13 |
| Generalisability | 21 | Discuss the generalisability (external validity) of the study results | 13 |
| **Other information** |  |  |  |
| Funding | 22 | Give the source of funding and the role of the funders for the present study and, if applicable, for the original study on which the present article is based | 14 |

\*Give information separately for cases and controls in case-control studies and, if applicable, for exposed and unexposed groups in cohort and cross-sectional studies.
**Note:** An Explanation and Elaboration article discusses each checklist item and gives methodological background and published examples of transparent reporting. The STROBE checklist is best used in conjunction with this article (freely available on the Web sites of PLoS Medicine at http://www.plosmedicine.org/, Annals of Internal Medicine at http://www.annals.org/, and Epidemiology at http://www.epidem.com/). Information on the STROBE Initiative is available at www.strobe-statement.org.

## References

1. Greenberg SM, Vernooij MW, Cordonnier C, Viswanathan A, Al-Shahi Salman R, Warach S, et al. Cerebral microbleeds: A guide to detection and interpretation. Lancet Neurol. 2009;8:165–174

2. Charidimou A, Shams S, Romero JR, Ding J, Veltkamp R, Horstmann S, et al. Clinical significance of cerebral microbleeds on mri: A comprehensive meta-analysis of risk of intracerebral hemorrhage, ischemic stroke, mortality, and dementia in cohort studies (v1). Int J Stroke. 2018;13:454–468

3. Akoudad S, Portegies ML, Koudstaal PJ, Hofman A, van der Lugt A, Ikram MA, et al. Cerebral microbleeds are associated with an increased risk of stroke: The rotterdam study. Circulation. 2015;132:509–516

4. Xu CX, Xu H, Yi T, Yi XY, Ma JP. Cerebral microbleed burden in ischemic stroke patients on aspirin: Prospective cohort of intracranial hemorrhage. Front Neurol. 2021;12:742899

5. Soo Y, Zietz A, Yiu B, Mok VCT, Polymeris AA, Seiffge D, et al. Impact of cerebral microbleeds in stroke patients with atrial fibrillation. Ann Neurol. 2023;94:61–74

6. Wang M, Yang Y, Wang Y, Luan M, Xu L, Zhong M, et al. Dual antiplatelet therapy in acute ischaemic stroke with or without cerebral microbleeds. Eur J Neurosci. 2023;57:1197–1207

7. Derraz I, Cagnazzo F, Gaillard N, Morganti R, Dargazanli C, Ahmed R, et al. Microbleeds, cerebral hemorrhage, and functional outcome after endovascular thrombectomy. Neurology. 2021;96:e1724–e1731

8. Frid P, Drake M, Giese AK, Wasselius J, Schirmer MD, Donahue KL, et al. Detailed phenotyping of posterior vs. Anterior circulation ischemic stroke: A multi-center mri study. J Neurol. 2020;267:649–658

9. Gregoire SM, Chaudhary UJ, Brown MM, Yousry TA, Kallis C, Jager HR, et al. The microbleed anatomical rating scale (mars): Reliability of a tool to map brain microbleeds. Neurology. 2009;73:1759–1766

10. North american symptomatic carotid endarterectomy trial. Methods, patient characteristics, and progress. Stroke. 1991;22:711–720

11. Samuels OB, Joseph GJ, Lynn MJ, Smith HA, Chimowitz MI. A standardized method for measuring intracranial arterial stenosis. AJNR Am J Neuroradiol. 2000;21:643–646

12. Kim BJ, Lee EJ, Kwon SU, Park JH, Kim YJ, Hong KS, et al. Prevention of cardiovascular events in asian patients with ischaemic stroke at high risk of cerebral haemorrhage (picasso): A multicentre, randomised controlled trial. Lancet Neurol. 2018;17:509–518

13. Hakim A, Gallucci L, Sperber C, Rezny-Kasprzak B, Jager E, Meinel T, et al. The analysis of association between single features of small vessel disease and stroke outcome shows the independent impact of the number of microbleeds and presence of lacunes. Sci Rep. 2024;14:3402

14. Low A, Mak E, Rowe JB, Markus HS, O’Brien JT. Inflammation and cerebral small vessel disease: A systematic review. Ageing Res Rev. 2019;53:100916

15. Yakushiji Y, Wilson D, Ambler G, Charidimou A, Beiser A, van Buchem MA, et al. Distribution of cerebral microbleeds in the east and west: Individual participant meta-analysis. Neurology. 2019;92:e1086–e1097

16. Luo Q, Tang H, Xu X, Huang J, Wang P, He G, et al. The prevalence and risk factors of cerebral microbleeds: A community-based study in china. Ther Clin Risk Manag. 2021;17:165–171

17. Cholesterol Treatment Trialists C. Efficacy and safety of statin therapy in older people: A meta-analysis of individual participant data from 28 randomised controlled trials. Lancet. 2019;393:407–415

18. Dawson J, Bejot Y, Christensen LM, De Marchis GM, Dichgans M, Hagberg G, et al. European stroke organisation (eso) guideline on pharmacological interventions for long-term secondary prevention after ischaemic stroke or transient ischaemic attack. Eur Stroke J. 2022;7:I–II

19. Kleindorfer DO, Towfighi A, Chaturvedi S, Cockroft KM, Gutierrez J, Lombardi-Hill D, et al. 2021 guideline for the prevention of stroke in patients with stroke and transient ischemic attack: A guideline from the american heart association/american stroke association. Stroke. 2021;52:e364–e467

20. Zhao Y, Zhou Y, Zhou H, Gong X, Luo Z, Li J, et al. Low-density lipoprotein cholesterol, statin therapy, and cerebral microbleeds: The circle study. Neuroimage Clin. 2023;39:103502

21. Ma C, Gurol ME, Huang Z, Lichtenstein AH, Wang X, Wang Y, et al. Low-density lipoprotein cholesterol and risk of intracerebral hemorrhage: A prospective study. Neurology. 2019;93:e445–e457

22. Nezu T, Hosomi N, Kitagawa K, Nagai Y, Nakagawa Y, Aoki S, et al. Effect of statin on stroke recurrence prevention at different infarction locations: A post hoc analysis of the j-stars study. J Atheroscler Thromb. 2020;27:524–533

23. Liu JY, Zhou YJ, Zhai FF, Han F, Zhou LX, Ni J, et al. Cerebral microbleeds are associated with loss of white matter integrity. AJNR Am J Neuroradiol. 2020;41:1397–1404

24. Elkhatib THM, Elsaid AF, Al-Molla RM, Khamis MEM, Fahmi RM. Prevalence and associated risk factors of cerebral microbleeds in egyptian patients with acute ischemic stroke and atrial fibrillation. J Stroke Cerebrovasc Dis. 2020;29:104703

25. Lau KK, Wong YK, Teo KC, Chang RSK, Tse MY, Hoi CP, et al. Long-term prognostic implications of cerebral microbleeds in chinese patients with ischemic stroke. J Am Heart Assoc. 2017;6

26. Ghaznawi R, Geerlings MI, Jaarsma-Coes M, Hendrikse J, de Bresser J, Group UC-SS. Association of white matter hyperintensity markers on mri and long-term risk of mortality and ischemic stroke: The smart-mr study. Neurology. 2021;96:e2172–e2183

27. Shoamanesh A, Hart RG, Connolly SJ, Kasner SE, Smith EE, Marti-Fabregas J, et al. Microbleeds and the effect of anticoagulation in patients with embolic stroke of undetermined source: An exploratory analysis of the navigate esus randomized clinical trial. JAMA Neurol. 2021;78:11–20

28. Wilson D, Ambler G, Lee KJ, Lim JS, Shiozawa M, Koga M, et al. Cerebral microbleeds and stroke risk after ischaemic stroke or transient ischaemic attack: A pooled analysis of individual patient data from cohort studies. Lancet Neurol. 2019;18:653–665

29. Sommer P, Posekany A, Serles W, Marko M, Scharer S, Fertl E, et al. Is functional outcome different in posterior and anterior circulation stroke? Stroke. 2018;49:2728–2732

30. Tong X, Liao X, Pan Y, Cao Y, Wang C, Liu L, et al. Intravenous thrombolysis is more safe and effective for posterior circulation stroke: Data from the thrombolysis implementation and monitor of acute ischemic stroke in china (tims-china). Medicine (Baltimore). 2016;95:e3848

